# Behavioral, Demographic, and Clinical determinants of HIV Status in Zambian Women

**DOI:** 10.1101/2021.05.05.21256411

**Authors:** Debebe Gebreyohannes, Ji Shen, Kelley Sams

## Abstract

The rate of human immunodeficiency virus (HIV) infection shows a diminishing trend globally while increasing in intensity of mortality, morbidity, and burden of HIV in Sub-Saharan Africa. The intertwined behavioral, demographic, and clinical determinants fueled the incidence of infections in Zambian women. This study aimed to determine the association between demographic, behavioral, and clinical determinants with HIV serostatus in Zambian women. With the conceptual framework of the World Health Organization’s Commission for Social Determinants of Health (CSDH) and the quantitative method of MANOVA, this study examined Zambian Demographic Health Survey data for Zambian women of two ages groups (adolescent and adult). The findings showed statistically significant results in the association between HIV serostatus and self-perceived HIV risk for both groups and in the association between education and HIV serostatus among women in both groups. However, there was no statistically significant association between behavioral, demographic, and clinical determinants of HIV serostatus. These findings imply the need to conduct prospective studies on such determinants to curb HIV and improve women’s community health in Africa.

**Author Summary:** Zambia is a country in the Sub-Saharan region of Africa, which is disproportionately facing the risk of HIV increase in infection rate and the number of people impacted. HIV exposure shows an extraordinary rise for women aged 18–49 years old. The behavioral, demographic, and clinical determinants of HIV serostatus form an intricate web that snares adolescent and adult women, deteriorating their quality of life and their mental and emotional well-being.

## Introduction

Human immunodeficiency virus (HIV) is a virus that infects humans and leads to acquired immune deficiency syndrome (AIDS), a lethal disease that weakens the immune system [1]. Approximately 37 million people live with the human immunodeficiency virus (HIV); about 2 million new infections and almost 1 million AIDS-related deaths occur each year [2].

Although infection rates of HIV/AIDS are decreasing globally, both infection rates and prevalence have increased in Zambia and other Sub-Saharan African countries [3, 4]. Meager HIV prevention and insufficient access to clinical services are common issues in countries in this region. Ninety percent of HIV infections in this region is due to low condom use and multiple sexual partners [5]. In Zambia, about 14% of people aged 18–49 years have HIV [6].

The purpose of this study was to assess the associations between the behavioral, demographic, and clinical determinants of HIV serostatus in Zambian women. Findings from this study may inform methods to reduce new infections, deter mortality and morbidity associated with AIDS, and improve the quality of life for Zambian women living with HIV/AIDS.

### Behavioral Factors

Behavioral factors, such as multiple sexual partners, unprotected sex, nondisclosure of HIV positive serostatus, increased HIV new infections, and spread of HIV for adolescent and adult women contributes to the prevalence of HIV [7, 8]. Mathur et al. found that, for heterosexual couples, having multiple sexual partners alone contributed to15% of female HIV-positive cases and increased HIV prevalence by 4% for 15- to 19-year-old Zambian women [9]. Other researchers have found a positive correlation between the rate of HIV infection and sexual risk-taking in adolescent and adult women [10]. For 15- to 49-year-olds, risky sexual behaviors led to more significant increases in HIV prevalence for women than for men [11].

### Demographic Factors

HIV serostatus correlates with demographic variables such as age, sex, and location. For example, Chanda-Kapata, Klinkenberg, Maddox, Ngosa, & Kapata [12] and McCarraher et al. [13] found that women have higher HIV infection rates than men. Furthermore, Okawa et al. [22] found that female adolescents did not understand HIV transmission methods, even though they were concerned about its effect in their future marriages. Kharsany and Karim [14] found consistent increases in HIV infection and transmission rates in Zambian women aged 18–49 years; these rates are six times higher than those of similarly aged Zambian men. Both HIV/AIDS infection rates and HIV-associated deaths are higher among women [15]. Research also suggests that the risk level is higher in urban areas than in rural areas [12,13]. People who are unaware of their HIV status are at an elevated risk of spreading the virus. In Zambia, HIV affects 2.9% of the population, and an estimated 1.2 million women aged 15–49 years have HIV [16].

### Clinical Factors

Clinical factors include HIV diagnosis, individuals’ relationships with their health care providers, and access to counseling. Some factors serve as barriers to HIV testing and treatment, whereas others (such as access to counseling) facilitate disease prevention.

## Materials and Methods

### Ethical Procedures

Ethical procedures were the formal requirements for conducting this study. I obtained a permission and an approval from Walden University Institutional Review Board before accessing and analyzing the the data for this study. The data description indicated the data were anonymous and personal information identifications were excluded to protect the anonymity of study participants.

### Population

The study population included Zambian women 18 to 49 years old who were permanent residents in 8 provinces. The target population was from a demographic health survey conducted between 2009 and 2010. The demographic health survey contained 1,4441 responses from Zambian women aged 18 to 49 years old about fertility preferences, HIV status; fertility preferences; attitudes toward and knowledge about HIV, and attitudes toward and use of HIV services [17].

### Sampling and Sampling Procedures

The sampling strategy divided each of the nine Zambian provinces Zambia into enumerative equal areas, based on the 2000 Zambian population and housing census. All households had the chance to be selected as part of the Zambian Demographic Health Survey (DHS) sample. The sampling procedure stratified the ZDHS sample into urban and rural areas. There were 18 sampling strata from the nine provinces. The samplers began with the Census Supervisory Area (CSA), then moved to the EA level, and finally independently selected enumeration areas for all stratifications. The sampling frame included Zambian women age 18 to 49 years old. The sampling frame excluded females younger than 18 years old, women older than 49 years old, and all males of any age. The research sample was sub-sampled from the 2007 Zambia Demographic and Health Survey (ZDHS). The sample was designed to provide specific indicators, including reproductive health indicators and HIV prevalence for each of the nine provinces in Zambia. Sorting the sample frame followed an implicit stratification and proportional allocation based on the geographical/administrative order and a probability proportional to size. Stratification sampling was completed before the selection of the sample [18]. Specific procedures guided the sample selection. Households were listed from 319 EAs. From each EA, an average of 25 households was selected through equal probability systematic sampling. Out of the average total of those EAs, the ones in Northern province and Lusaka province were sub-sampled that shall be used for this study. I used a power analysis to determine the sample size. The power analysis to determine the sample size included the justification for the effect size, alpha level, the power level, and the citation source for calculating the sample size.

## Results

The purpose of the study was to investigate the association between dependent variables (HIV serostatus) and independent variables (behavioral determinants, demographic determinants, and clinical determinants) for Zambian women aged 18–49 years using secondary data collected in 2009–2010 from Zambia demographic health survey. In addition, I compared adolescents (aged 18–24 years) to adults (25–49 years) to determine whether there were differences in the associations between behavioral, demographic, and clinical determinants and that of the synergistic association of behavioral, demographic, and clinical determinants and HIV serostatus between these age groups.

A multiple analysis of variance (MANOVA) demonstrated a statistically significant (*p*< 0.01) for the association between self-perceived HIV risk and HIV serostatus for Zambian women aged 18 – 24 (see *Table 1*).

**Table 1.**
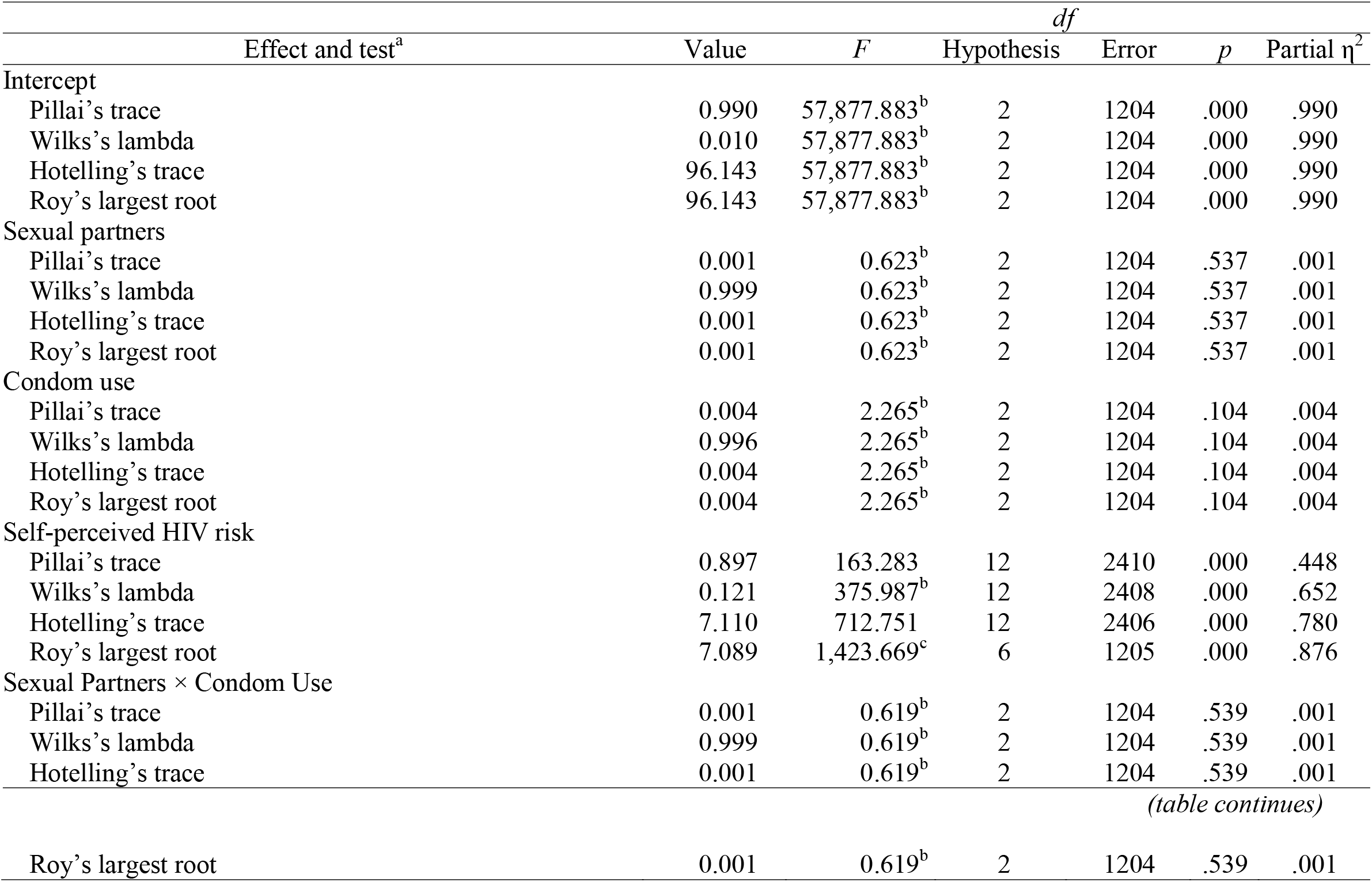
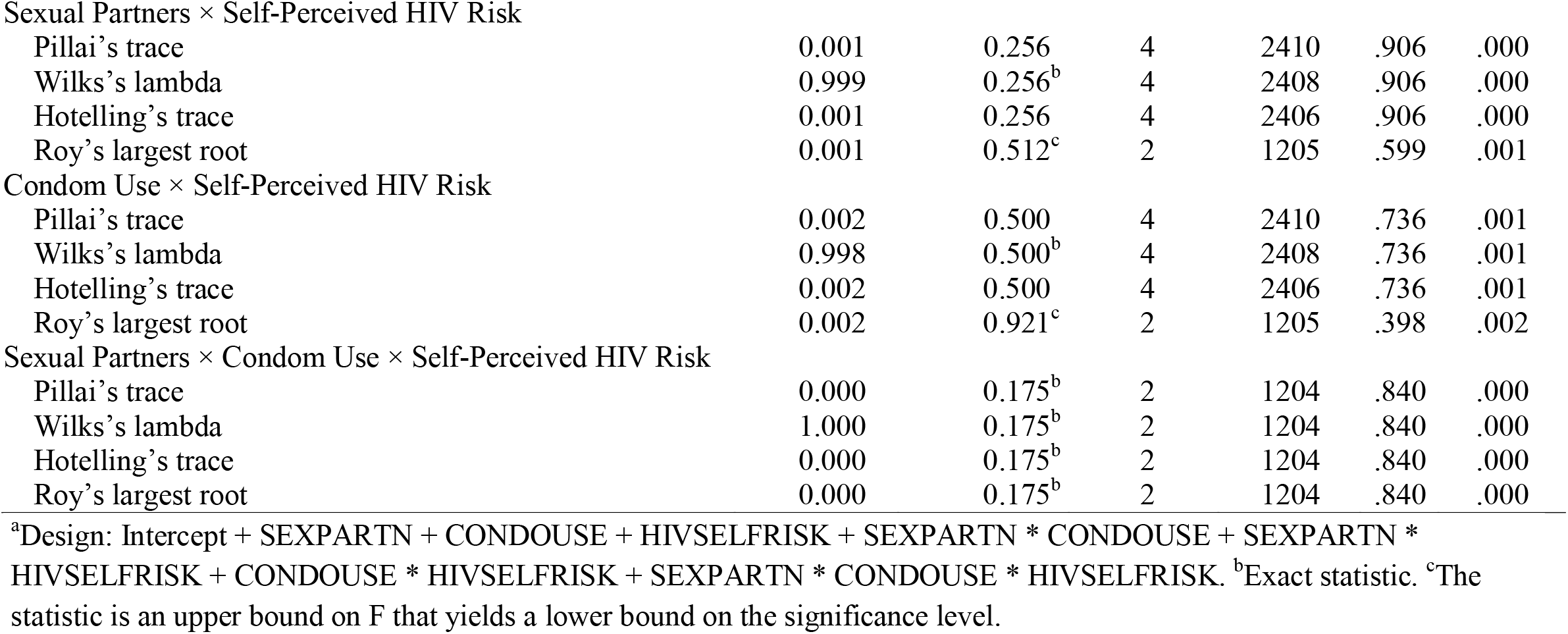
Multivariate Tests on Behavioral Determinants of HIV Serostatus for Zambian Women Aged 18–24 Years

The multivariate tests for associations between the dependent variables: HIV test result, ever tested HIV, and ever tested AIDS) and behavioral determinants: sexual partners, condom use, and self-perceived risk for women aged 25–49. There was a statistically significant difference between self-perceived HIV risk and a linear combination of the three dependent variables. In this main effect, self-perceived HIV risk was statistically significant: Pillai’s trace was 0.897, F(2, 2410) = 163.283, p < .001; Wilks’s lambda = 0.121, partial η2 = .448 (see *Table 2*).

**Table 2.**
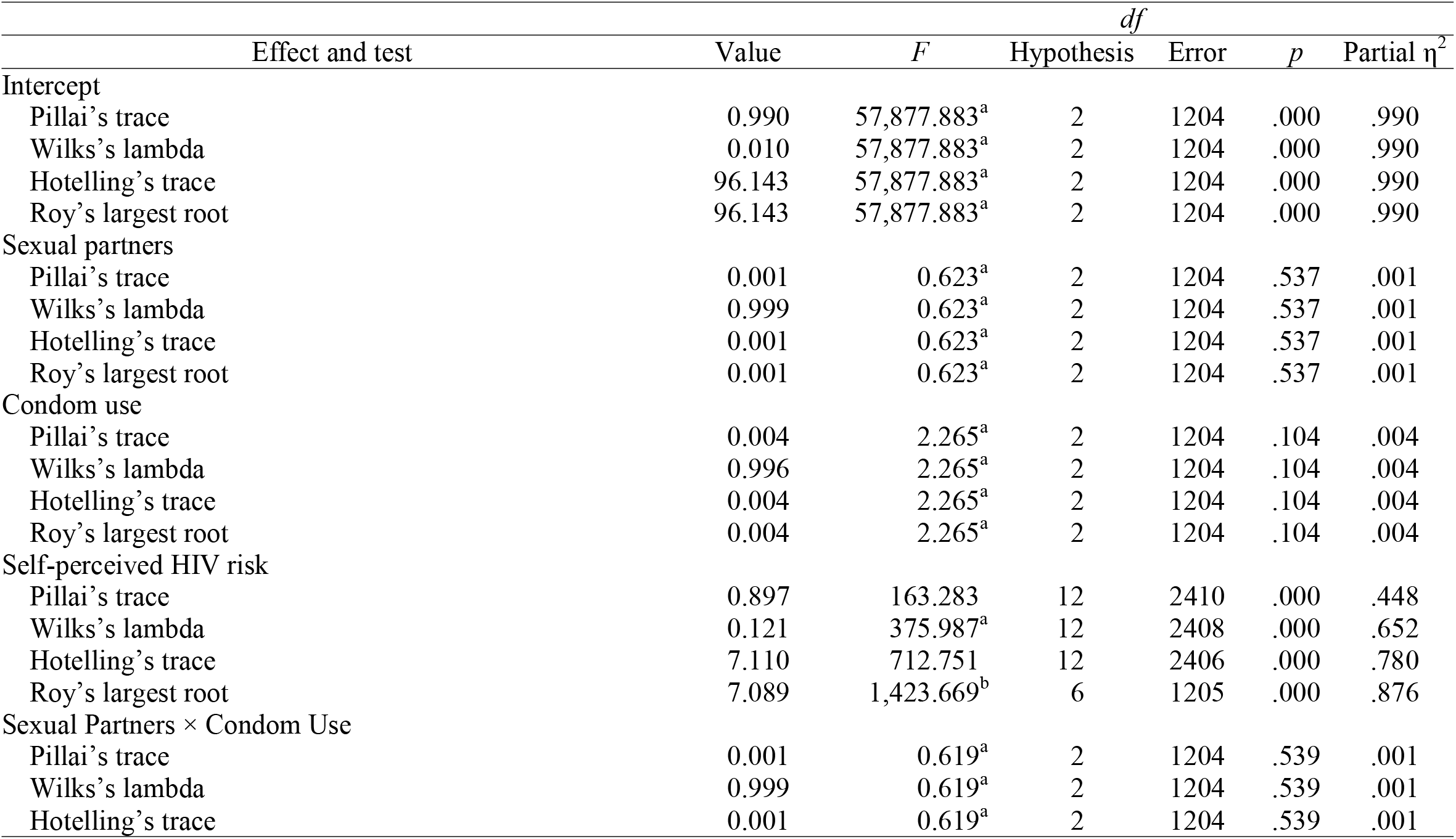
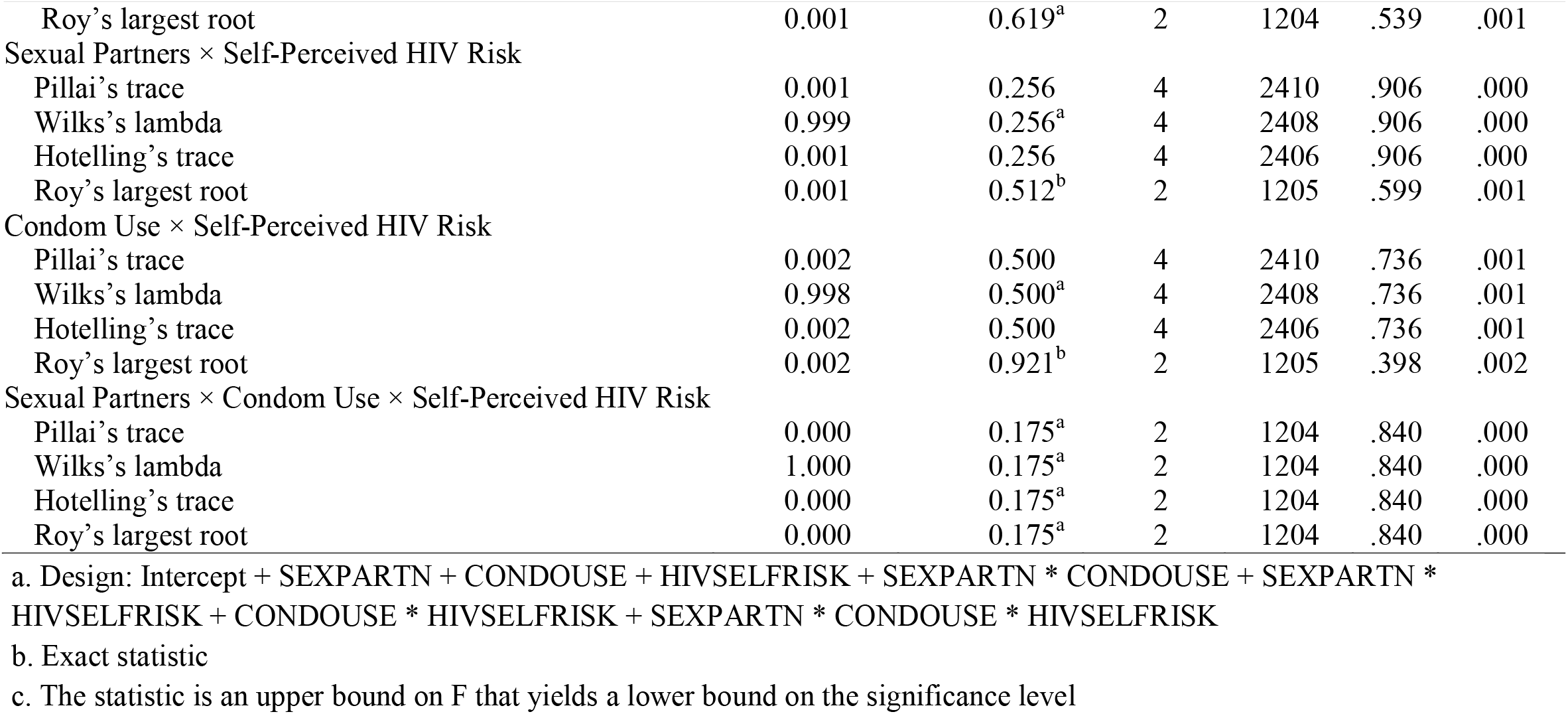
Multivariate Tests on Behavioral Determinants of HIV Serostatus on Zambian Women Aged 25–49 Years

The multivariate test demonstrated as follows: there was no statistically significant association between demographic determinants and HIV serostatus of Zambian female adolescents from ages 18 to 24. Tables 10 and 11 indicate the MANOVA results of hypothesis testing 3 by analyzing the associations between demographic determinants (location, education, and marital status) with the dependent variable HIV serostatus. It appeared a statistically significant association between education and a linear combination of the three dependent variables (see *Table 3*).

**Table 3.** Multivariate Tests on Demographic Determinants of HIV Serostatus on Zambian Women Aged 18–24 Years

| Effect and test<sup>a</sup> | Value | *F* | Hypothesis | *df* | | *p* | Partial $\eta^2$ |
| --- | --- | --- | --- | --- | --- | --- | --- |
|  |  |  |  | Error |  |  |  |
| Intercept |  |  |  |  |  |  |  |
| Pillai's trace | 0.927 | 5,109.357<sup>b</sup> | 3 | 1215.000 |  | .000 | .927 |
| Wilks's lambda | 0.073 | 5,109.357<sup>b</sup> | 3 | 1215.000 |  | .000 | .927 |
| Hotelling's trace | 12.616 | 5,109.357<sup>b</sup> | 3 | 1215.000 |  | .000 | .927 |
| Roy's largest root | 12.616 | 5,109.357<sup>b</sup> | 3 | 1215.000 |  | .000 | .927 |
| Location |  |  |  |  |  |  |  |
| Pillai's trace | 0.002 | 0.914<sup>b</sup> | 3 | 1215.000 |  | .433 | .002 |
| Wilks's lambda | 0.998 | 0.914<sup>b</sup> | 3 | 1215.000 |  | .433 | .002 |
| Hotelling's trace | 0.002 | 0.914<sup>b</sup> | 3 | 1215.000 |  | .433 | .002 |
| Roy's largest root | 0.002 | 0.914<sup>b</sup> | 3 | 1215.000 |  | .433 | .002 |
| Education |  |  |  |  |  |  |  |
| Pillai's trace | 0.015 | 2.099 | 9 | 3651.000 |  | .026 | .005 |
| Wilks's lambda | 0.985 | 2.106 | 9 | 2957.141 |  | .026 | .005 |
| Hotelling's trace | 0.016 | 2.111 | 9 | 3641.000 |  | .025 | .005 |
| Roy's largest root | 0.015 | 5.930<sup>c</sup> | 3 | 1217.000 |  | .001 | .014 |
| Marital status |  |  |  |  |  |  |  |
| Pillai's trace | 0.003 | 0.363 | 9 | 3651.000 |  | .953 | .001 |
| Wilks's lambda | 0.997 | 0.363 | 9 | 2957.141 |  | .953 | .001 |
| Hotelling's trace | 0.003 | 0.362 | 9 | 3641.000 |  | .953 | .001 |
| Roy's largest root | 0.002 | 0.964<sup>c</sup> | 3 | 1217.000 |  | .409 | .002 |
| Location × Education |  |  |  |  |  |  |  |
| Pillai's trace | 0.002 | 0.225 | 9 | 3651.000 |  | .991 | .001 |
| Wilks's lambda | 0.998 | 0.224 | 9 | 2957.141 |  | .991 | .001 |
| Hotelling's trace | 0.002 | 0.224 | 9 | 3641.000 |  | .991 | .001 |

*(table continues)*

| Effect and test<sup>a</sup> | Value | *F* | *df* | | *p* | Partial $\eta^2$ |
| --- | --- | --- | --- | --- | --- | --- |
|  |  |  | Hypothesis | Error |  |  |
| Roy's largest root | 0.001 | 0.496<sup>c</sup> | 3 | 1217.000 | .685 | .001 |
| Location × Marital Status | | | | | | |
| Pillai's trace | 0.004 | 0.588 | 9 | 3651.000 | .808 | .001 |
| Wilks's lambda | 0.996 | 0.588 | 9 | 2957.141 | .808 | .001 |
| Hotelling's trace | 0.004 | 0.588 | 9 | 3641.000 | .808 | .001 |
| Roy's largest root | 0.004 | 1.576<sup>c</sup> | 3 | 1217.000 | .193 | .004 |
| Education × Marital Status | | | | | | |
| Pillai's trace | 0.012 | 0.587 | 24 | 3651.000 | .944 | .004 |
| Wilks's lambda | 0.988 | 0.587 | 24 | 3524.471 | .944 | .004 |
| Hotelling's trace | 0.012 | 0.588 | 24 | 3641.000 | .944 | .004 |
| Roy's largest root | 0.010 | 1.449<sup>c</sup> | 8 | 1217.000 | .172 | .009 |
| Location × Education × Marital Status | | | | | | |
| Pillai's trace | 0.007 | 0.506 | 18 | 3651.000 | .957 | .002 |
| Wilks's lambda | 0.993 | 0.506 | 18 | 3437.024 | .957 | .002 |
| Hotelling's trace | 0.007 | 0.505 | 18 | 3641.000 | .957 | .002 |
| Roy's largest root | 0.005 | 1.039<sup>c</sup> | 6 | 1217.000 | .398 | .005 |

a. Design: Intercept + LOCATION + EDUCATION + MARITALST + LOCATION \* EDUCATION + LOCATION \* MARITALST + EDUCATION \* MARITALST + LOCATION \* EDUCATION \* MARITALST

Multivariate Tests on demographic determinants of HIV Serostatus on Zambian Women Aged 25–49 years indicated a statistically significant association between education and HIV serostatus for women aged 25–49 years. In this main effect education was statistically significant in its association with HIV serostatus: Pillai’s trace was 0.015, F(9, 3651) = 2.099, p = .026; Wilks’s lambda was 0.985, partial η2 = .005 (see *Table 4*).

**Table 4.**
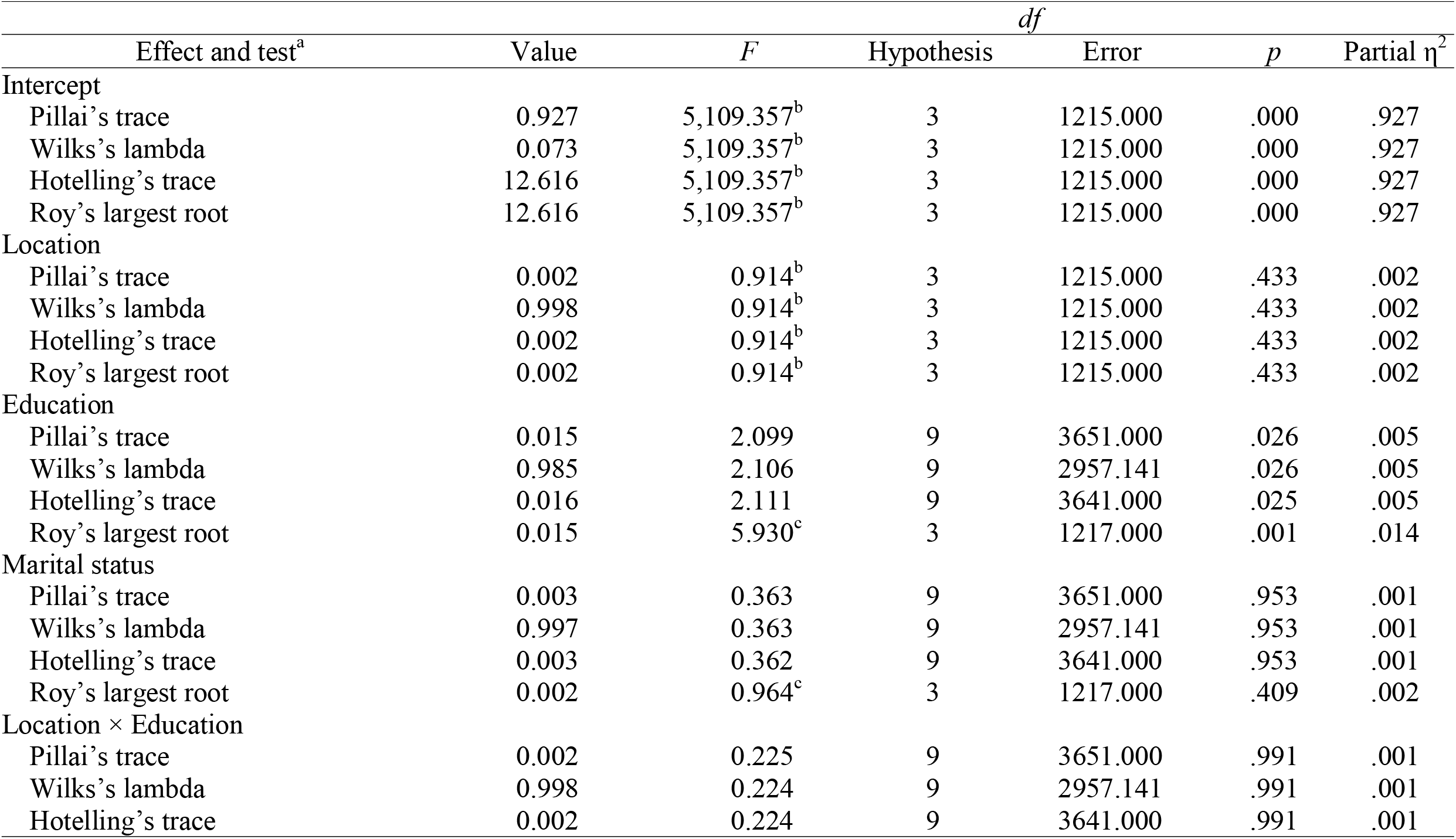
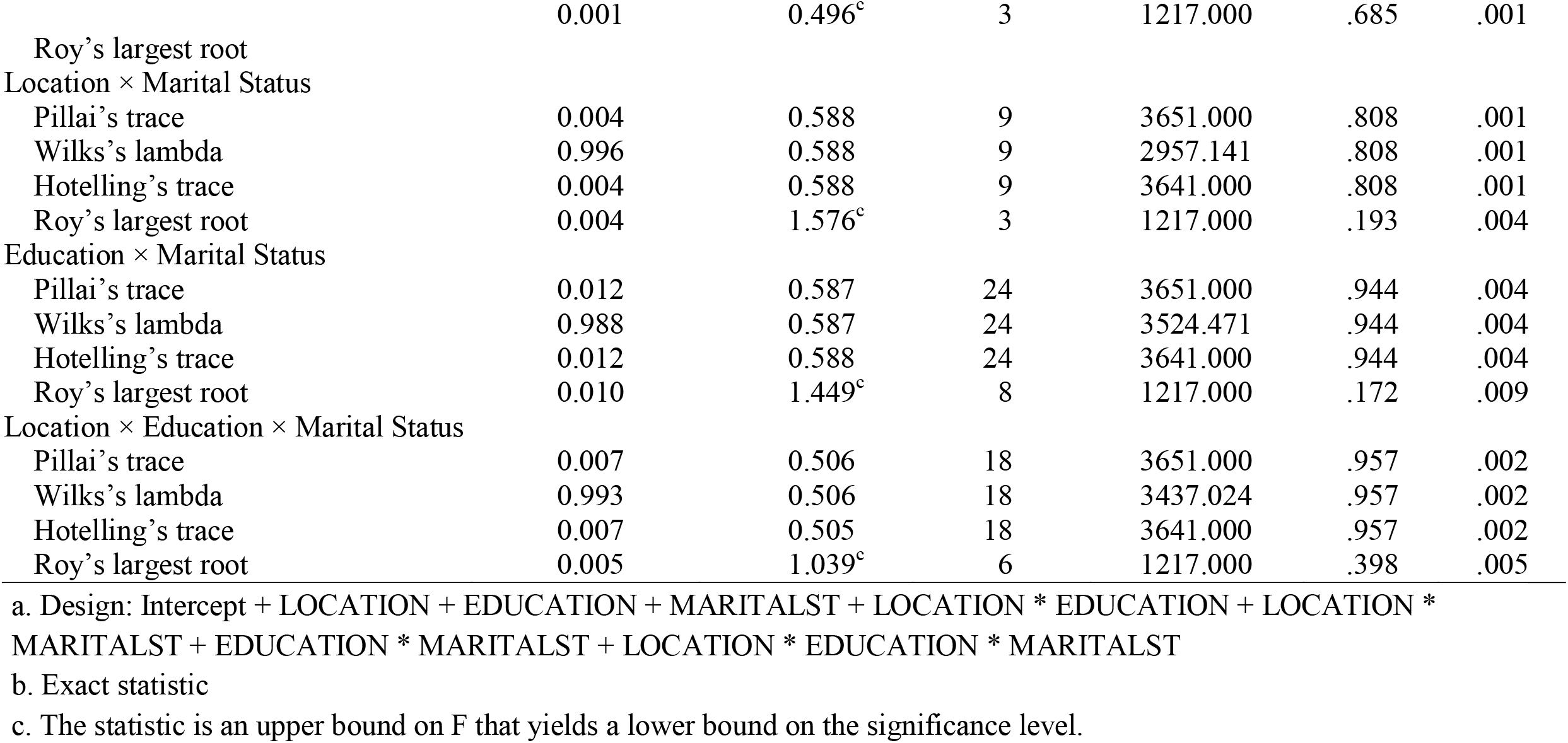
Multivariate Tests on Demographic Determinants of HIV Serostatus on Zambian Women Aged 25–49 Years

The multivariate tests showed no statistically significant associations between clinical determinants (government services, clinic services, NGO services) with HIV serostatus for Zambian women of 25 - 49 years (see *Table 5*).

**Table 5.** Multivariate Tests on Clinical Determinants of HIV Serostatus for Zambian Women Aged 18–24 Years

| Effect and test<sup>a</sup> | Value | $F^b$ | $p$ | Partial $\eta^2$ |
| --- | --- | --- | --- | --- |
| Intercept |  |  |  |  |
| Pillai's trace | 0.986 | 43,506.206 | .000 | .986 |
| Wilks's lambda | 0.014 | 43,506.206 | .000 | .986 |
| Hotelling's trace | 71.380 | 43,506.206 | .000 | .986 |
| Roy's largest root | 71.380 | 43,506.206 | .000 | .986 |
| Clinic services |  |  |  |  |
| Pillai's trace | 0.000 | 0.170 | .844 | .000 |
| Wilks's lambda | 1.000 | 0.170 | .844 | .000 |
| Hotelling's trace | 0.000 | 0.170 | .844 | .000 |
| Roy's largest root | 0.000 | 0.170 | .844 | .000 |
| NGO Services |  |  |  |  |
| Pillai's trace | 0.003 | 1.658 | .191 | .003 |
| Wilks's lambda | 0.997 | 1.658 | .191 | .003 |
| Hotelling's trace | 0.003 | 1.658 | .191 | .003 |
| Roy's largest root | 0.003 | 1.658 | .191 | .003 |
| Government services |  |  |  |  |
| Pillai's trace | 0.000 | 0.287 | .751 | .000 |
| Wilks's lambda | 1.000 | 0.287 | .751 | .000 |
| Hotelling's trace | 0.000 | 0.287 | .751 | .000 |
| Roy's largest root | 0.000 | 0.287 | .751 | .000 |
| Clinic Services × NGO Services | | | | |
| Pillai's trace | 0.000 | 0.002 | .998 | .000 |
| Wilks's lambda | 1.000 | 0.002 | .998 | .000 |
| Hotelling's trace | 0.000 | 0.002 | .998 | .000 |
| Roy's largest root | 0.000 | 0.002 | .998 | .000 |

*(table continues)*

| Effect and test<sup>a</sup> | Value | $F^b$ | $p$ | Partial $\eta^2$ |
| --- | --- | --- | --- | --- |
| Clinic Services × Government Services | | | | |
| Pillai's trace | 0.000 | 0.006 | .994 | .000 |
| Wilks's lambda | 1.000 | 0.006 | .994 | .000 |
| Hotelling's trace | 0.000 | 0.006 | .994 | .000 |
| Roy's largest root | 0.000 | 0.006 | .994 | .000 |
| NGO Services × Government Services | | | | |
| Pillai's trace | 0.001 | 0.597 | .551 | .001 |
| Wilks's lambda | 0.999 | 0.597 | .551 | .001 |
| Hotelling's trace | 0.001 | 0.597 | .551 | .001 |
| Roy's largest root | 0.001 | 0.597 | .551 | .001 |
| Clinic Services × NGO Services × Government Services | | | | |
| Pillai's trace | 0.000 | 0.089 | .915 | .000 |
| Wilks's lambda | 1.000 | 0.089 | .915 | .000 |
| Hotelling's trace | 0.000 | 0.089 | .915 | .000 |
| Roy's largest root | 0.000 | 0.089 | .915 | .000 |

*Note.* Hypothesis $df = 2$ . Error $df = 1219$ . NGO = nongovernmental organization.
a. Design: Intercept + GOVTEST + CLINICTEST + NGOTEST + GOVTEST \* CLINICTEST + GOVTEST \* NGOTEST + CLINICTEST \* NGOTEST + GOVTEST \* CLINICTEST \* NGOTEST

As shown below, there were no statistically significant differences between clinical determinants and a linear combination of the three dependent variables (HIV serostatus, ever tested HIV, ever tested AIDS) for adult Zambian women of 25 – 49 years (see *Table 6*).

**Table 6.**
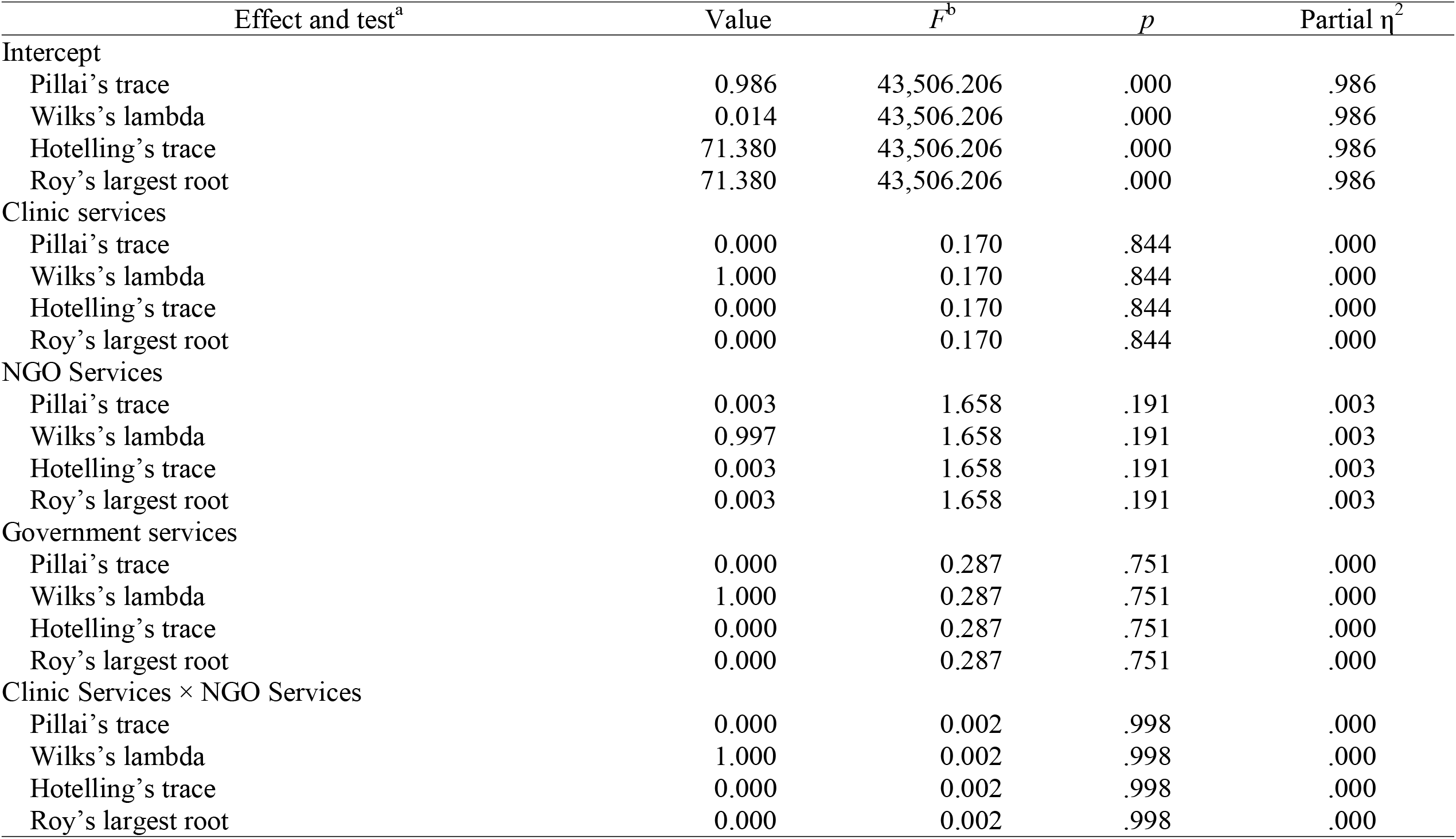
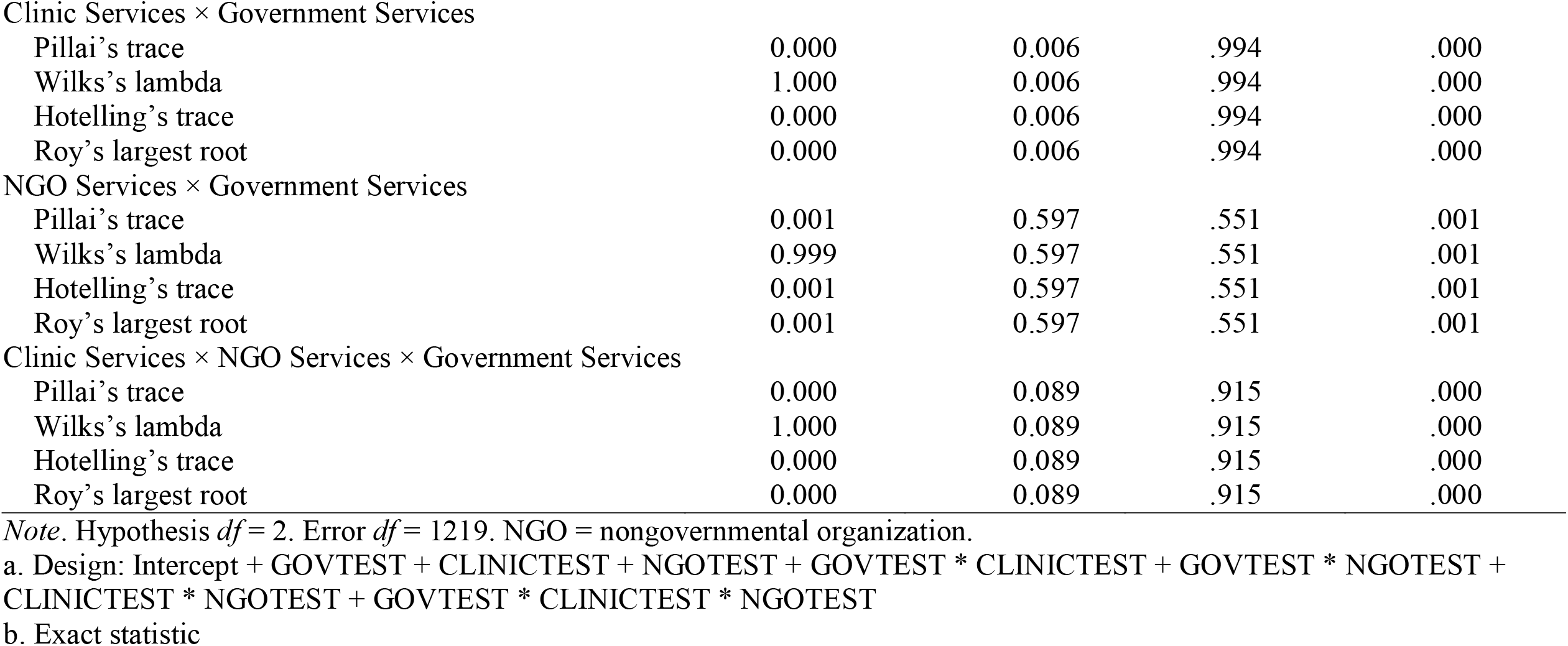
Multivariate Tests on Clinical Determinants of HIV Serostatus for Zambian Women Aged 25–49 Years

Multivariate analysis of variance was also performed to test hypothesis 7 stated as follows: there is no statistically significant synergistic association between behavioral, demographic, and clinical determinants and that of HIV serostatus of Zambian female adolescents from age 18 to 24. In addition, the synergistic associations between sexual partners, location, and government with HIV test results were not statistically significant (*Table 7*).

**Table 7.** Multivariate Tests on Synergistic Analysis of Behavioral, Demographic, and Clinical Determinants of HIV Serostatus for Zambian Women Aged 18–24 Years

| Effect and test<sup>a</sup> | Value | *F* | Hypothesis | *df* | | *p* | Partial $\eta^2$ |
| --- | --- | --- | --- | --- | --- | --- | --- |
|  |  |  |  | Error |  |  |  |
| Intercept |  |  |  |  |  |  |  |
| Pillai's trace | 0.991 | 63,188.837<sup>b</sup> | 2 | 1195.0 |  | .000 | .991 |
| Wilks's lambda | 0.009 | 63,188.837<sup>b</sup> | 2 | 1195.0 |  | .000 | .991 |
| Hotelling's trace | 105.755 | 63,188.837<sup>b</sup> | 2 | 1195.0 |  | .000 | .991 |
| Roy's largest root | 105.755 | 63,188.837<sup>b</sup> | 2 | 1195.0 |  | .000 | .991 |
| Sexual partners |  |  |  |  |  |  |  |
| Pillai's trace | 0.001 | 0.257 | 4 | 2392.0 |  | .905 | .000 |
| Wilks's lambda | 0.999 | 0.257<sup>b</sup> | 4 | 2390.0 |  | .905 | .000 |
| Hotelling's trace | 0.001 | 0.257 | 4 | 2388.0 |  | .905 | .000 |
| Roy's largest root | 0.001 | 0.508<sup>c</sup> | 2 | 1196.0 |  | .602 | .001 |
| Location |  |  |  |  |  |  |  |
| Pillai's trace | 0.001 | 0.438<sup>b</sup> | 2 | 1195.0 |  | .645 | .001 |
| Wilks's lambda | 0.999 | 0.438<sup>b</sup> | 2 | 1195.0 |  | .645 | .001 |
| Hotelling's trace | 0.001 | 0.438<sup>b</sup> | 2 | 1195.0 |  | .645 | .001 |
| Roy's largest root | 0.001 | 0.438<sup>b</sup> | 2 | 1195.0 |  | .645 | .001 |
| Government services |  |  |  |  |  |  |  |
| Pillai's trace | 0.870 | 460.457 | 4 | 2392.0 |  | .000 | .435 |
| Wilks's lambda | 0.130 | 1,058.488<sup>b</sup> | 4 | 2390.0 |  | .000 | .639 |
| Hotelling's trace | 6.680 | 1,993.840 | 4 | 2388.0 |  | .000 | .770 |
| Roy's largest root | 6.679 | 3,994.196<sup>c</sup> | 2 | 1196.0 |  | .000 | .870 |
| Sexual Partners × Location | | | | | | | |
| Pillai's trace | 0.004 | 1.285 | 4 | 2392.0 |  | .273 | .002 |
| *(table continues)* |  |  |  |  |  |  |  |
| Wilks's lambda | 0.996 | 1.285<sup>b</sup> | 4 | 2390.0 |  | .273 | .002 |

| Effect and test<sup>a</sup> | Value | *F* | *df* | | *p* | Partial $\eta^2$ |
| --- | --- | --- | --- | --- | --- | --- |
|  |  |  | Hypothesis | Error |  |  |
| Hotelling's trace | 0.004 | 1.285 | 4 | 2388.0 | .273 | .002 |
| Roy's largest root | 0.004 | 2.471<sup>c</sup> | 2 | 1196.0 | .085 | .004 |
| Sexual Partners × Government Services | | | | | | |
| Pillai's trace | 0.001 | 0.416<sup>b</sup> | 2 | 1195.0 | .660 | .001 |
| Wilks's lambda | 0.999 | 0.416<sup>b</sup> | 2 | 1195.0 | .660 | .001 |
| Hotelling's trace | 0.001 | 0.416<sup>b</sup> | 2 | 1195.0 | .660 | .001 |
| Roy's largest root | 0.001 | 0.416<sup>b</sup> | 2 | 1195.0 | .660 | .001 |
| Location × Government Services | | | | | | |
| Pillai's trace | 0.000 | 0.039 | 4 | 2392.0 | .997 | .000 |
| Wilks's lambda | 1.000 | 0.038<sup>b</sup> | 4 | 2390.0 | .997 | .000 |
| Hotelling's trace | 0.000 | 0.038 | 4 | 2388.0 | .997 | .000 |
| Roy's largest root | 0.000 | 0.077<sup>c</sup> | 2 | 1196.0 | .926 | .000 |
| Sexual Partners × Location × Government Services | | | | | | |
| Pillai's trace | 0.000 | —<sup>b</sup> | 0 | 0.0 | — | — |
| Wilks's lambda | 1.000 | —<sup>b</sup> | 0 | 1195.5 | — | — |
| Hotelling's trace | 0.000 | —<sup>b</sup> | 0 | 2.0 | — | — |
| Roy's largest root | 0.000 | 0.000<sup>b</sup> | 2 | 1194.0 | 1.000 | .000 |

a. Design: Intercept + SEXPARTN + LOCATION + GOVTEST + SEXPARTN \* LOCATION + SEXPARTN \* GOVTEST + LOCATION \* GOVTEST + SEXPARTN \* LOCATION \* GOVTEST
b. Exact statistic
c. The statistic is an upper bound on *F* that yields a lower bound on the significance level.

Multivariate analysis of variance (MANOVA) revealed no statistically significant synergistic association between behavioral, demographic, and clinical determinants and the HIV test result in Zambian female adults from age 25 to 49 (see *Table 8*).

**Table 8.** Multivariate Tests on Synergistic Association of Behavioral, Demographic, and Clinical Determinants of HIV Serostatus for Zambian Women Aged 25–49 Years

| Effect and test<sup>a</sup> | Value | *F* | Hypothesis | *df* | | *p* | Partial $\eta^2$ |
| --- | --- | --- | --- | --- | --- | --- | --- |
|  |  |  |  | Error |  |  |  |
| Intercept |  |  |  |  |  |  |  |
| Pillai's trace | 0.987 | 43575.177<sup>b</sup> | 2 | 1183 |  | .000 | .987 |
| Wilks's lambda | 0.013 | 43575.177<sup>b</sup> | 2 | 1183 |  | .000 | .987 |
| Hotelling's trace | 73.669 | 43575.177<sup>b</sup> | 2 | 1183 |  | .000 | .987 |
| Roy's largest root | 73.669 | 43575.177<sup>b</sup> | 2 | 1183 |  | .000 | .987 |
| Self-perceived HIV risk |  |  |  |  |  |  |  |
| Pillai's trace | 0.011 | 1.292 | 10 | 2368 |  | .229 | .005 |
| Wilks's lambda | 0.989 | 1.294<sup>b</sup> | 10 | 2366 |  | .228 | .005 |
| Hotelling's trace | 0.011 | 1.297 | 10 | 2364 |  | .226 | .005 |
| Roy's largest root | 0.011 | 2.597<sup>c</sup> | 5 | 1184 |  | .024 | .011 |
| Marital status |  |  |  |  |  |  |  |
| Pillai's trace | 0.000 | 0.051 | 6 | 2368 |  | .999 | .000 |
| Wilks's lambda | 1.000 | 0.051<sup>b</sup> | 6 | 2366 |  | .999 | .000 |
| Hotelling's trace | 0.000 | 0.051 | 6 | 2364 |  | .999 | .000 |
| Roy's largest root | 0.000 | 0.102<sup>c</sup> | 3 | 1184 |  | .959 | .000 |
| NGO Services |  |  |  |  |  |  |  |
| Pillai's trace | 0.001 | 0.552<sup>b</sup> | 2 | 1183 |  | .576 | .001 |
| Wilks's lambda | 0.999 | 0.552<sup>b</sup> | 2 | 1183 |  | .576 | .001 |
| Hotelling's trace | 0.001 | 0.552<sup>b</sup> | 2 | 1183 |  | .576 | .001 |
| Roy's largest root | 0.001 | 0.552<sup>b</sup> | 2 | 1183 |  | .576 | .001 |
| Self-Perceived HIV Risk × Marital Status | | | | | | | |
| Pillai's trace | 0.008 | 0.329 | 28 | 2368 |  | 1.000 | .004 |
| Wilks's lambda | 0.992 | 0.329<sup>b</sup> | 28 | 2366 |  | 1.000 | .004 |

| Effect and test<sup>a</sup> | Value | *F* | *df* | | *p* | Partial $\eta^2$ |
| --- | --- | --- | --- | --- | --- | --- |
|  |  |  | Hypothesis | Error |  |  |
| *(table continues)* |  |  |  |  |  |  |
| Hotelling's trace | 0.008 | 0.329 | 28 | 2364 | 1.000 | .004 |
| Roy's largest root | 0.008 | 0.657<sup>c</sup> | 14 | 1184 | .817 | .008 |
| Self-Perceived HIV Risk × NGO Services | | | | | | |
| Pillai's trace | 0.002 | 0.186 | 10 | 2368 | .997 | .001 |
| Wilks's lambda | 0.998 | 0.186<sup>b</sup> | 10 | 2366 | .997 | .001 |
| Hotelling's trace | 0.002 | 0.186 | 10 | 2364 | .997 | .001 |
| Roy's largest root | 0.002 | 0.370<sup>c</sup> | 5 | 1184 | .869 | .002 |
| Marital Status × NGO Services | | | | | | |
| Pillai's trace | 0.002 | 0.333 | 6 | 2368 | .920 | .001 |
| Wilks's lambda | 0.998 | 0.333<sup>b</sup> | 6 | 2366 | .920 | .001 |
| Hotelling's trace | 0.002 | 0.333 | 6 | 2364 | .920 | .001 |
| Roy's largest root | 0.002 | 0.665<sup>c</sup> | 3 | 1184 | .574 | .002 |
| Self-Perceived HIV Risk × Marital Status × NGO Services | | | | | | |
| Pillai's trace | 0.002 | 0.264 | 10 | 2368 | .989 | .001 |
| Wilks's lambda | 0.998 | 0.264<sup>b</sup> | 10 | 2366 | .989 | .001 |
| Hotelling's trace | 0.002 | 0.264 | 10 | 2364 | .989 | .001 |
| Roy's largest root | 0.002 | 0.527<sup>c</sup> | 5 | 1184 | .756 | .002 |

*Note.* NGO = nongovernmental organization.
a. Design: Intercept + CONDOUSE + EDUCATION + CLINICTEST + CONDOUSE \* EDUCATION + CONDOUSE \* CLINICTEST + EDUCATION \* CLINICTEST + CONDOUSE \* EDUCATION \* CLINICTEST
b. Exact statistic
c. The statistic is an upper bound on *F* that yields a lower bound on the significance level.

## Discussion

### Behavioral Determinants of HIV Serostatus

For both women aged 18–24 years and women aged 25–49 years, there was a significant association between HIV serostatus and self-perceived HIV risk but not sexual partners or condom use. This finding is partly consistent with Adeniyi et al. [19], who also examined behavioral, demographic, and clinical determinants and HIV serostatus. However, Adeniyi et al.[19] studied only one HIV serostatus variable compared to this study’s three serostatus variables.

The findings indicated a link between the HIV test result variable and self-perceived HIV risk for women of18–24 years. There was a strong association between the perception of no risk or small risk and HIV serostatus. Toska et al.[10] reported the presence of a direct association between HIV acquisition of adolescent women (aged 18–24 years) and having multiple sexual partners, although the finding was inconclusive. That finding was in harmony with a secondary analysis of a cross-sectional study conducted in Uganda, which indicated an association between self-perceived HIV risk and incidence of HIV infection among women of all ages. This study elucidates the impact of self-perceived HIV risk. It suggests a prevention strategy for Zambian women and also probably women in similar settings and socioeconomic statuses in other parts of sub-Saharan Africa. Rosenberg et al. [20], reporting on a population-based study, said that the spread of HIV had accelerated in older adults in South Africa and recommended the commencement of prevention activities for analogous communities in sub-Saharan African countries.

### Demographic Determinants of HIV Serostatus

The findings indicated a statistically significant association between education and HIV serostatus among women in both age groups. However, unlike previous studies, there were no statistically significant differences for the demographic variables of location and marital status and pairwise interactions of location and marital status, location and education, and marital status and education. Omonaiye, Kusljic, Nicholson, and Manias [21], Pinchoff, Boyer, Mutombo, Chowdhuri, and Ngo [7], and Okawa et al. [22] reported that age and education were critical to determining HIV serostatus for adolescent and adult women at individual and societal levels.

Lack of associations between HIV serostatus and age (adolescent versus adult) and location (urban versus rural) also contrasted with the findings of authors who stated strong and significant associations between HIV serostatus and age, gender, and location [12,13]. Mee et al. [23] found that being in school increased students’ awareness of HIV and the practice of preventive measures and eventually contributed to diminishing the spread of HIV infection.

Other researchers failed to find an association between education and the use of HIV services [24]. Bunyasi and Coetzee [25] suggested a direct association between accomplishment in education and the reduction of HIV prevalence. The main reason for the association between a lower incidence of HIV and higher educational achievement may be the ability of educated people to implement safer sexual practices and abstain from sexual activity. Increased health literacy and academic progress could empower educated people to make informed decisions and take responsibility for themselves and their communities.

### Clinical Determinants of HIV Serostatus

The findings indicated a significant association between aggregate use of HIV services and HIV serostatus for women in both age groups. This result neither confirmed nor refuted the conclusions reported by authors of systematic reviews, who implied that it was essential to conduct further studies to maximize prevention through the use of HIV medications [26,27]. However, there were no significant associations between HIV serostatus and the use of government services, clinic services, and NGO services considered individually. This finding agreed with the results of Mustapha, Musiime, Bakeera-Kitaka, Rujumba, and Nabukeera-Barungi [28] regarding the use of HIV services by women in Uganda. Those authors stated that HIV services for women were substandard in one sub-Saharan country, Uganda. Mustapha et al. [28] reported that women received motivation to use services from knowing HIV status and demotivation to use services from not knowing HIV serostatus, discrimination, and economic constraints.

Authors of systematic studies described the benefits of team-based HIV service access to improve HIV prevention in sub-Saharan African countries. Reviewers of quantitative and qualitative studies explicitly described associations between HIV serostatus and the use of team-based services versus institutionalized HIV services—government, clinic, and NGO [29]. The availability of HIV services in HIV-prone sub-Saharan African countries could be positive indicators of the containment of HIV spread and the deterrence of new infection. The use of HIV services in communities has the same level of importance. Along with HIV services, the implementation of precise HIV diagnostic tests to minimize false test outcomes is critical in sub-Saharan African countries [30].

### Demographic, Behavioral, and Clinical Determinants of HIV Serostatus

Multivariate tests indicated no significant association between the synergistic association of Demographic, Behavioral, and Clinical Determinants with HIV Serostatus for Zambian women aged 18–24. Likewise, there was no significant association between sub-Saharan Africa [31]. The finding regarding the synergistic association between behavioral, demographic, and clinical determinants with HIV serostatus could not address the recommendation by Kharsany and Karim [14] for a study of behavioral determinants and the HIV literacy (education) gap concerning HIV serostatus cognizance at the community level. Synergistic association of Demographic, Behavioral, and Clinical Determinants with HIV Serostatus for Zambian women aged 25 to 49. The community strategy outlined by health professionals, policymakers, and other entities involved in HIV prevention should be a collective drive to manage HIV epidemics.

### Limitations of the Study

This study had certain limitations. The exclusion of those aged younger than 18 years and older than 49 years, prisoners, and military personnel was one of the limitations. Because resource constraints dictated that the ZDHS surveyed only two provinces, the other eight provinces were excluded from data collection. The complete absence of men could be a significant limitation. Another limitation was that the study was cross-sectional rather than longitudinal. Thus, it lacked the benefits of conducting longitudinal research, including identifying temporal effects and associated changes in the dynamics of sexual behaviors, demographic shifts, and capacity and acceptability of HIV prevention services. The last limitation was the age of the data from the ZDHS. The laps of 10 years since the data were collected means that the data may not reflect the current situation. However, I could overcome this to some extent by extrapolating the findings through the lens of other consistent HIV research findings in sub-Saharan Africa. Thus, generalization of the results beyond the specific setting to other similar settings appears reasonable.

## Recommendations

The recommendations in this section apply to Zambian women and women living in similar settings throughout sub-Saharan Africa. The consequences and risks of HIV prevail in older women. Future researchers can improve the robustness of their findings by including older women. Including this age group in HIV prevention efforts and will assist in significantly diminishing HIV in sub-Saharan Africa. According to Maughan-Brown and Venkataramani [32]), the high prevalence of HIV in South Africa was because of risky sexual behaviors among older women. National and international collaboration are pivotal to lessen the spread of HIV throughout the world because HIV is a public health threat in every country. A unified approach to HIV prevention and control would have collateral health benefits when supported with measurable goals and comprehensive strategies [33,34].

### Implications

The dynamics of sexual behaviors and their associations with HIV serostatus indicate their impact on HIV prevalence, incidence, and epidemics in sub-Saharan Africa. One of the focuses was the paramount importance of the consolidation of leadership to mobilize communities for harmonized HIV response [35]. In conjunction with the findings on the associations of behavioral, demographic, and clinical determinants with HIV serostatus for Zambian women aged 18–49, the inclusion of female participants younger than 18 years and older than 49 years would yield more holistic findings that could be applied to fighting HIV [20].

## Conclusions

The findings regarding the associations of behavioral, demographic, and clinical determinants with HIV serostatus can be used to project risk, inform prevention efforts, and initiate longitudinal research. These findings suggest that self-perceived HIV risk, education, and type of HIV services are reliable indicators of HIV serostatus. Effective HIV prevention and control in sub-Saharan countries should focus on developing Awareness using educational advancement as a tool, recognizing HIV risks through health literacy, and addressing social determinants of HIV health. These findings complement growing evidence indicating the benefits of collaborative HIV prevention efforts that involve HIV researchers and health professionals at both the local and international levels.

## Data Availability

Publicly available at ICPSR under the title "Survey of HIV Status and Fertility Preferences in Sub-Saharan Africa, 2009-2010 (ICPSR 36718)"
Bankole, Akinrinola

https://www.debebe.manuscriptone.com

## Acknowledgments

The corresponding author is grateful to Dr. Ji Shen and Dr. Kelly Sams, who supported me with their expertise to complete my dissertation research at Walden University. I extend the incredible support of Dr. Ji Shen and Dr. Kelly Sams to editing this manuscript to submit to Plos One Publishing Agency. I believe they will remain to be part of my academic journey to publish future scholarly articles.

## References

1. CDC. CDC Works 24/7 [Internet]. Cdc.gov. 2021 [cited 2021 Feb 8]. Available from: http://www.cdc.gov/

2. Schwetz TA, Fauci AS. The extended impact of human immunodeficiency virus/active immunodeficiency syndrome research. Journal of Infectious Diseases. 2019;219(1):6–9.

3. Ranganathan M, MacPhail C, Pettifor A, Kahn K, Khoza N, Twine R, et al. Young women’s perceptions of transactional sex and sexual agency: A qualitative study in the context of rural South Africa. BMC Public Health. 2017;17:1–16.

4. Wakeham K, Harding R, Levin J, Parkes-Ratanshi R, Kamali A, Lalloo DG. The impact of antiretroviral therapy on symptom burden among human immunodeficiency virus outpatients with low cluster of differentiation 4 (CD4) count in rural Uganda: Nested longitudinal cohort study. BMC Palliative Care. 2017;17:1–9.

5. Muzyamba C, Broaddus E, Campbell C. “You cannot eat rights”: a qualitative study of views by Zambian HIV-vulnerable women, youth and MSM on human rights as public health tools. BMC International Health and Human Rights. 2015 Dec;15(1):1–6.

6. Amoyaw JA, Kuuire VZ, Boateng GO, Asare-Bediako Y, Ung M. Conundrum of sexual decision making in marital relationships: safer-sex knowledge, behavior, and attitudes of married women in Zambia. The Journal of Sex Research. 2015 Oct 13;52(8):868–77.

7. Pinchoff J, Boyer CB, Mutombo N, Chowdhuri RN, Ngo TD. Why don’t urban youth in Zambia use condoms? The influence of gender and marriage on non-use of male condoms among young adults. PloS one. 2017 Mar 23;12(3):e0172062.

8. Salam RA, Faqqah A, Sajjad N, Lassi ZS, Das JK, Kaufman M, Bhutta ZA. Improving adolescent sexual and reproductive health: A systematic review of potential interventions. Journal of adolescent health. 2016 Oct 1;59(4): S11–28.

9. Mathur S, Okal J, Musheke M, Pilgrim N, Kishor Patel S, Bhattacharya R, et al. High rates of sexual violence by both intimate and non-intimate partners experienced by adolescents girls and young women in Kenya and Zambia: Findings around violence and other negative health outcomes. PLoS ONE. 2018 Sep 13;13(9):1–13.

10. Toska E, Pantelic M, Meinck F, Keck K, Haghighat R, Cluver L. Sex in the shadow of HIV: A systematic review of prevalence, risk factors, and interventions to reduce sexual risk-taking among HIV-positive adolescents and youth in sub-Saharan Africa. PLoS ONE. 2017 Jun 5 [cited 2021 Mar 12];12(6):1–30.

11. Hegdahl HK, Fylkesnes KM, Sandøy IF. Sex Differences in HIV Prevalence Persist over Time: Evidence from 18 Countries in Sub-Saharan Africa. PLoS ONE. 2016 Feb 3;11(2):1–17.

12. Chanda-Kapata P, Kapata N, Klinkenberg E, William N, Mazyanga L, Musukwa K, Kawesha EC, Masiye F, Mwaba P. The adult prevalence of HIV in Zambia: results from a population-based mobile testing survey conducted in 2013–2014. AIDS research and therapy. 2016 Dec;13(1):1–9. doi:10.1186/si2981-015-0088-1

13. McCarraher DR, Packer C, Mercer S, Dennis A, Banda H, Nyambe N, Stalter RM, Mwansa JK, Katayamoyo P, Denison JA. Adolescents living with HIV in the Copperbelt Province of Zambia: Their reproductive health needs and experiences. PloS one. 2018 Jun 5;13(6):e0197853. doi: 10.1371/journal.pone.0197853

14. Kharsany AB, Karim QA. HIV Infection and AIDS in Sub-Saharan Africa: Current Status, Challenges and Opportunities. Open AIDS J. 2016 Apr 8;10:34–48. doi: 10.2174/1874613601610010034.

15. Amoyaw JA, Kuuire VZ, Boateng GO, Asare-Bediako Y, Ung M. Conundrum of Sexual Decision Making in Marital Relationships: Safer-Sex Knowledge, Behavior, and Attitudes of Married Women in Zambia. Journal of Sex Research. 2015 Oct;52(8):868–77. doi: 10.1080/00224499.2014.996280

16. Qiao S, Zhang Y, Li X, Menon JA. Facilitators and barriers for HIV-testing in Zambia: A systematic review of multi-level factors. PLoS ONE. 2018 Feb 7;13(2):1–27.

17. Bankole, A. Survey of HIV Status and Fertility Preferences in Sub-Saharan Africa. Ann Arbor, MI: Inter-university Consortium for Political and Social Research [distributor], 2017-04-10. 2009-2010. doi.org/10.3886/ICPSR36718.v2

18. Creswell, J. W. (2014). Research design: Qualitative, quantitative, and mixed methods (4th ed.). Thousand Oaks, CA: Sage. 2014.

19. Adeniyi OV, Ajayi AI, Selanto-Chairman N, Goon DT, Boon G, Fuentes YO, Hofmeyr GJ, Avramovic G, Carty C, Lambert J, East London Prospective Cohort Study (ELPCS) Group. Demographic, clinical and behavioural determinants of HIV serostatus nondisclosure to sex partners among HIV-infected pregnant women in the Eastern Cape, South Africa. PloS one. 2017 Aug 24;12(8):e0181730. doi:10.1371/journal.pone.0181730

20. Rosenberg MS, Gómez-Olivé FX, Rohr JK, Houle BC, Kabudula CW, Wagner RG, et al. Sexual Behaviors and HIV Status: A Population-Based Study Among Older Adults in Rural South Africa. JAIDS: Journal of Acquired Immune Deficiency Syndromes. 2017 Jan;74(1):e9–17. doi:10.1097/QAI.0000000000001173

21. Omonaiye O, Kusljic S, Nicholson P, Manias E. Medication adherence in pregnant women with human immunodeficiency virus receiving antiretroviral therapy in sub-Saharan Africa: a systematic review. BMC Public Health. 2018 Dec;18(1):1–20. doi:10.1186/s12889-018-5651-y

22. Okawa S, Mwanza-Kabaghe S, Mwiya M, Kikuchi K, Jimba M, Kankasa C, Ishikawa N. Sexual and reproductive health behavior and unmet needs among a sample of adolescents living with HIV in Zambia: a cross-sectional study. Reproductive health. 2018 Dec;15(1):1–0. doi:10.1186/s12978-018-0493-8

23. Mee P, Fearon E, Hassan S, Hensen B, Acharya X, Rice BD, Hargreaves JR. The association between being currently in school and HIV prevalence among young women in nine eastern and southern African countries. PloS one. 2018 Jun 20;13(6):e0198898.

24. Sabapathy K., Hensen B., Varsaneux O., Floyd S., Fidler S., Hayes R. The cascade of care following community-based detection of human immunodeficiency virus in sub-Saharan Africa—a systematic review with 90-90-90 targets in sight. PLOS One, Published: July 27, 2018 https://doi.org/10.1371/journal.pone.0200737

25. Bunyasi EW, Coetzee D.J. Relationship between socioeconomic status and HIV infection: findings from a survey in the Free State and Western Cape Provinces of South Africa. BMJ open. 2017 Nov 1;7(11).

26. Mark D, Armstrong A, Andrade C, Penazzato M, Hatane L, Taing L, Runciman T, Ferguson J. HIV treatment and care services for adolescents: a situational analysis of 218 facilities in 23 sub□JSaharan African countries. Journal of the International AIDS Society. 2017 May;20:21591.

27. Williams S, Renju J, Ghilardi L, Wringe A. Scaling a waterfall: a meta-ethnography of adolescent progression through the stages of HIV care in sub-Saharan Africa. Journal of the International AIDS Society. 2017 Sep 15; 20:1–17.

28. Mustapha M, Musiime V, Bakeera-Kitaka S, Rujumba J, Nabukeera-Barungi N. Utilization of “prevention of mother-to-child transmission” of HIV services by adolescent and young mothers in Mulago Hospital, Uganda. BMC Infectious Diseases [Internet]. 2018 Nov 14;18(1).:N.PAG.

29. Mukumbang FC, Van Belle S, Marchal B, van Wyk B. An exploration of group-based HIV/AIDS treatment and care models in Sub-Saharan Africa using a realist evaluation (Intervention-Context-Actor-Mechanism-Outcome) heuristic tool: a systematic review. Implementation Science. 2017 Aug 252:1–20.

30. Kravitz D. S., Parekh B., Douglas M.O., Edgil D, Kuritsky J, Nkengasong J. A Commitment to HIV Diagnostic Accuracy–a comment on “Towards more accurate HIV testing in sub□Saharan Africa: a multi□site evaluation of HIV RDT s and risk factors for false positives” and “HIV misdiagnosis in sub□Saharan Africa: a performance of diagnostic algorithms at six testing sites”. Journal of the International AIDS Society. 2018 Aug;21(8):e25177.

31. Bibiana NE, Emmanuel PO, Amos D, Ramsey YM, Idris AN. Knowledge, attitude and factors affecting voluntary HIV counseling and testing services among women of reproductive age group in an Abuja Suburb community, Nigeria. Medical Journal of Zambia. 2018;45(1):13–22.

32. Maughan-Brown B, Venkataramani AS. Accuracy and determinants of perceived HIV risk among young women in South Africa. BMC public health. 2018 Dec;18(1):1–9.

33. Baron D, Essien T, Pato S, Magongo M, Mbandazayo N, Scorgie F, Rees H, Delany□Moretlwe S. Collateral benefits: how the practical application of Good Participatory Practice can strengthen HIV research in sub□Saharan Africa. Journal of the International AIDS Society. 2018 Oct;21:e25175.

34. Jones J, Sullivan PS, Curran JW. Progress in the HIV epidemic: Identifying goals and measuring success. PLoS medicine. 2019 Jan 18;16(1):e1002729.

35. Fakoya AO, Dybul MR, Sands P. Getting local: focusing on communities to achieve greater impact in the next phase of the HIV response. Journal of the International AIDS Society. 2019 Jul;22(7).

